# Single center blind testing of a US multi-center validated diagnostic algorithm for Kawasaki disease in Asia

**DOI:** 10.1101/2022.05.12.22275028

**Authors:** Ho-Chang Kuo, Shiying Hao, Bo Jin, C. James Chou, Zhi Han, Ling-Sai Chang, Ying-Hsien Huang, KuoYuan Hwa, John C. Whitin, Karl G. Sylvester, Charitha D. Reddy, Henry Chubb, Scott R. Ceresnak, John T. Kanegaye, Adriana H. Tremoulet, Jane C. Burns, Doff McElhinney, Harvey J. Cohen, Xuefeng B. Ling

## Abstract

Kawasaki disease (KD) is the leading cause of acquired heart disease in children. A key objective of research in KD is to reduce the risk of long-term cardiovascular sequelae by expediting timely diagnosis. The major challenge in KD diagnosis is that it shares clinical signs with other childhood febrile illnesses. Our previously single-center developed computer-based two-step algorithm was further tested by a five-center validation in US. This first blinded multi-center trial validated our approach, with sufficient sensitivity and positive predictive value, to identify most patients with KD diagnosed at centers across the US.

We sought to determine if our algorithmic approach applied to an Asian cohort. This study involved 418 KD and 259 febrile controls (FC) from the Chang Gung Memorial Hospital in Taiwan. Our diagnostic algorithm retained sensitivity (379 of 418; 90.7%), specificity (223 of 259; 86.1%), PPV (379 of 409; 92.7%), and NPV (223 of 247; 90.3%) comparable to previous US 2016 single center and US 2020 fiver center results. Only 4.7% (15 of 418) of KD and 2.3% (6 of 259) of FC patients were identified as indeterminate. The algorithm identified 18 of 50 (36%) KD patients who presented 2 or 3 principal criteria. Of 418 KD patients, 157 were infants younger than one year and 89.2% (140 of 157) were classified correctly. Of the 44 patients with KD who had coronary artery abnormalities, our diagnostic algorithm correctly identified 43 (97.7%) including all patients with dilated coronary artery but one who found to resolve in 8 weeks.

We assessed the performance of our KD diagnostic algorithm with a single center Asian cohort. This work demonstrates the applicability of our algorithmic approach and diagnostic portability, providing evidence to support the launch of an adequately powered, multicenter study for future Asian application in the emergency department setting. If deployed in Asia, our tool promises a cost-effective diagnostic approach to allow the timely management of Asian KD patients even in the absence of KD experts, to potentially enhance the outcome for KD patients and reduce the risk of coronary artery aneurysms.

## INTRODUCTION

Kawasaki disease (KD) is an acute vasculitis that affects infants and children and is the leading cause of acquired pediatric heart disease in the U.S. and Japan [1]. Timely and accurately diagnosis of KD is critical, yet there isn’t a gold standard diagnostic test for KD. The classic KD diagnostic criteria [2, 3] adopted by the American Heart Association (AHA) include fever plus ≥4 of 5 principal clinical signs, which include rash, conjunctival injection, extremity changes, oropharyngeal changes, and swollen lymph nodes. However, KD shares these clinical signs with other childhood febrile illnesses, causing missed or delayed diagnosis in emergency departments (EDs).

Among patients ultimately diagnosed with KD, only 4.7% receive the correct diagnosis at the first medical visit [4]. Delayed diagnosis causes delayed treatment [5] and thus increases the risk of developing aneurysm, and in some children leads to myocardial infarction or death [6]. Delayed diagnosis is particularly problematic among patients with incomplete clinical manifestations of KD (having less than 4 principal clinical signs), despite supplementary laboratory criteria have been adopted by AHA to identify atypical KD cases. Furthermore, treatment with intravenous immunoglobulin (IVIG), which can reduce the incidence of coronary aneurysms and risk of long-term cardiovascular complications [2, 7], is recommended to be given within 10 days of illness. There is a critical need for clinicians to timely and accurately differentiate KD from other pediatric febrile illnesses.

We have explored to apply an algorithmic approach to the bedside electronic health record (EHR) datasets, developing predictive analytics [8-37] to drive translational medicine for improved diagnosis of high impact diseases and prediction of clinical resource utilization. Regarding KD, in 2013, we tested the hypothesis whether statistical learning on clinical and laboratory test patterns can lead to a single-step algorithm for KD diagnosis [10]. Combining both clinical and laboratory test results, the algorithm diagnosed with sensitivity of 74.3% and specificity of 62.8% with > 95% PPV and > 95% NPV. In 2016, we improved and validated a two-step algorithm to classify an individual as a patient with KD, a febrile control (FC) or intermediate [21]. In the single-center validation, the algorithm yielded a sensitivity of 96.0% and a specificity of 78.5% with > 95% PPV and > 95% NPV. We subsequently set to validate this two-step diagnostic algorithm with five pediatric hospitals in the USA [31]: Boston Children’s Hospital, Boston, Massachusetts; Children’s Hospital Colorado, Aurora, Colorado; Children’s Hospital of Orange County, Orange, California; Nationwide Children’s Hospital, Columbus, Ohio; and Rady Children’s Hospital, San Diego, California. The blinded US multicenter validation validates the algorithm with a sensitivity of 91.6%, specificity of 57.8% and PPV and NPV of 95.4% and 93.1%, respectively. The algorithm also correctly identified 94.0% of KD patients with abnormal echocardiograms.

In this study, we performed a blinded, single center validation of this KD diagnostic algorithm in Taiwan. Validating the algorithmic portability from US to a single center in Asia, we set to evaluate this algorithm in a prospective multi-center trial in Asia.

## MATERIAL AND METHODS

### Study design

We previously developed and validated, with data from a US single center, a two-step algorithm that applies a linear discriminant analysis (LDA)-based model followed by a random forest-based algorithm to differentiate patients with KD from children with other febrile controls (FCs) [8, 10, 21, 31]. Patients who are classified as indeterminate by the LDA-based model are then evaluated by the random-forest algorithm based on the number of KD clinical criteria with which they present. In line with previous approach [8, 10, 21, 31], we applied the two-step algorithm, constructed with a single-center US cohort (Rady Children’s Hospital, San Diego, California, the validation cohort in 2016 study [21]), to a single center in Taiwan. The evaluation was blinded to diagnosis during classification, and then unblinded to calculate performance.

### Study population

A single center (Chang Gung Memorial Hospital in Taiwan) cohort of patients with acute KD were prospectively enrolled by local KD specialists. Signed consent or assent forms were obtained from the parents of all subjects and from all subjects >6 years of age. The study was approved by the institutional review boards of the Chang Gung Memorial Hospital and Stanford University. The diagnosis of KD was made by Taiwan KD specialists on the 2004 USA AHA guidelines [3]. All FCs had unexplained fever, ≥1 of the five principal clinical criteria for KD, and laboratory evaluation. All FC diagnosis was determined by clinical features, culture or PCR testing. All patients had fever (≥38.0° C) for no more than 10 days and had complete information for nine clinical or laboratory data points, including the five principal clinical criteria (illness days (ie, days of fever), total white cell count (WCC), percentage of eosinophils and hemoglobin concentration), percentages of monocytes, lymphocytes, neutrophils and immature neutrophils (bands), platelet count, levels of C reactive protein, (CRP), gamma glutamyl transferase (GGT), alanine aminotransferase (ALT) and erythrocyte sedimentation rate (ESR), gamma glutamyl transferase (GGT), were collected if available. The clinical parameters of bands, GGT, and ESR were not routinely collected in the Taiwan Chang Gung Hospital for the KD and FC patients, therefore, not used in the two-step algorithm. Age and sex for all patients, and coronary artery status and Z-score (SD from the mean adjusted for body surface area) for patients with KD, were also recorded when available. Coronary artery status was classified as normal (right coronary artery (RCA) and left anterior descending (LAD) Z-score always <2.5) or abnormal (RCA and/or LAD Z-score ≥2.5 within the first 6weeks after diagnosis).

### Two-step algorithm for differentiation of KD and FCs

The two-step algorithm uses an LDA-based analysis followed by a random forest-based algorithm as previously described [8, 21, 31]. In this study, input variables of the LDA model are illness days, the 5 principal clinical criteria and 10 laboratory test variables. The variables and their corresponding weights in the algorithm are listed in Supplementary Table 1. One or more variables had missing values for 227 of 418 (54.3%) patients with KD and 256 of 259 (98.8%) FCs. The algorithm was less sensitive in identifying KD patients with 1 or more missing variables as compared to patients with complete data. Values were imputed with a k-nearest neighbor algorithm. The output of the LDA classifies each individual patient as KD, FC or indeterminate. The cut-off thresholds between the three categories were set in the original US single center algorithm to provide a positive predictive value (PPV) or negative positive value (NPV) of 95%. The indeterminate patients were divided into four sub cohorts based on the number of KD criteria present at the time of diagnosis, and separate random forest models were applied to the patients in each sub-cohort to further stratify these patients into KD, FC or indeterminate categories [10, 21, 31].

**Table 1.** Demographic characteristics of the testing cohort in Taiwan.

| Characteristics | Taiwan Single Center Cohort |  |  |
| --- | --- | --- | --- |
|  | KD (n=418) | FC (n=259) | *p*-Value |
| Age, years,<br>median (IQR<sup>a</sup>) | 1.3<br>(0.7, 2.4) | 1.0<br>(0.6, 4.6) | 0.03 |
| Males, n (%) | 253 (60.5) | 138 (53.3) | 0.06 |
| Number of KD<br>criteria met, n (%) |  |  |  |
| ≤3 | 50 (12.0) | 217 (83.8) |  |
| 4 or 5 | 368 (88.0) | 42 (16.2) |  |

### Algorithm performance in this Asia single center study

The aim of this study was to analyze the algorithm performance on a single center Taiwan cohort. We compared the classifications of patients by the algorithm with the classifications made by the clinical experts who diagnosed these patients. We calculated sensitivity, specificity, PPV, NPV and rates of indeterminate classification of the algorithm when applied to this cohort. Performance of the algorithm was evaluated according to age, illness days and coronary artery status. We also performed univariable analysis with each of the 15 variables in sub cohorts of patients manifesting ⩾2, 3 or ≥4 KD principal clinical criteria. In each sub cohort, the distribution of each variable was compared between the patients with KD and FCs, and the differences were measured using Mann-Whitney U test (for continuous variables) or Fisher’s exact test (for categorical variables).

## RESULTS

### Patient characteristics

As shown in Figure 1, 751 patients were enrolled with 462 KD and 289 FC subjects. 26 FC patients were excluded due to incorrect registration of days of fever. 47 patients (KD: 44; FC: 3) were excluded with days of fever greater than 10 days. One FC patient was excluded due to the absence of KD principal criteria. For this study, from the Taiwan single center cohort (Table 1), 418 patients with KD and 259 FCs were used for the blind testing. Patients with KD were more likely to be male than FCs (60.5% vs 53.3%, *p*=0.06). The KDs had a higher median age than the patients with FC (1.3 vs 1.0 years, *p*=0.03). The FCs most common final diagnosis was either bacterial or viral infection (Table 2).

**Table 2.**
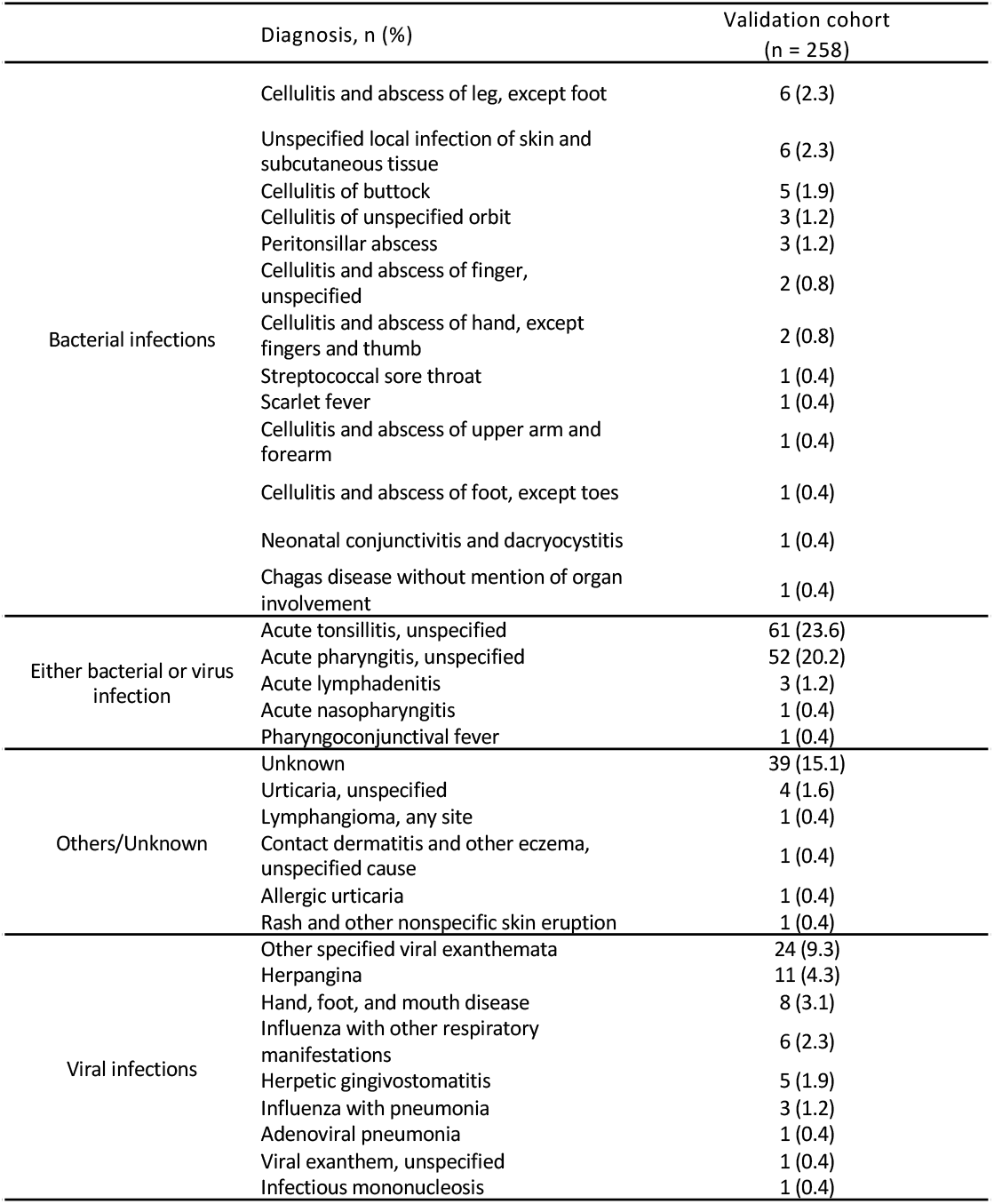
The final diagnosis of the febrile illness controls.

**Figure 1.**
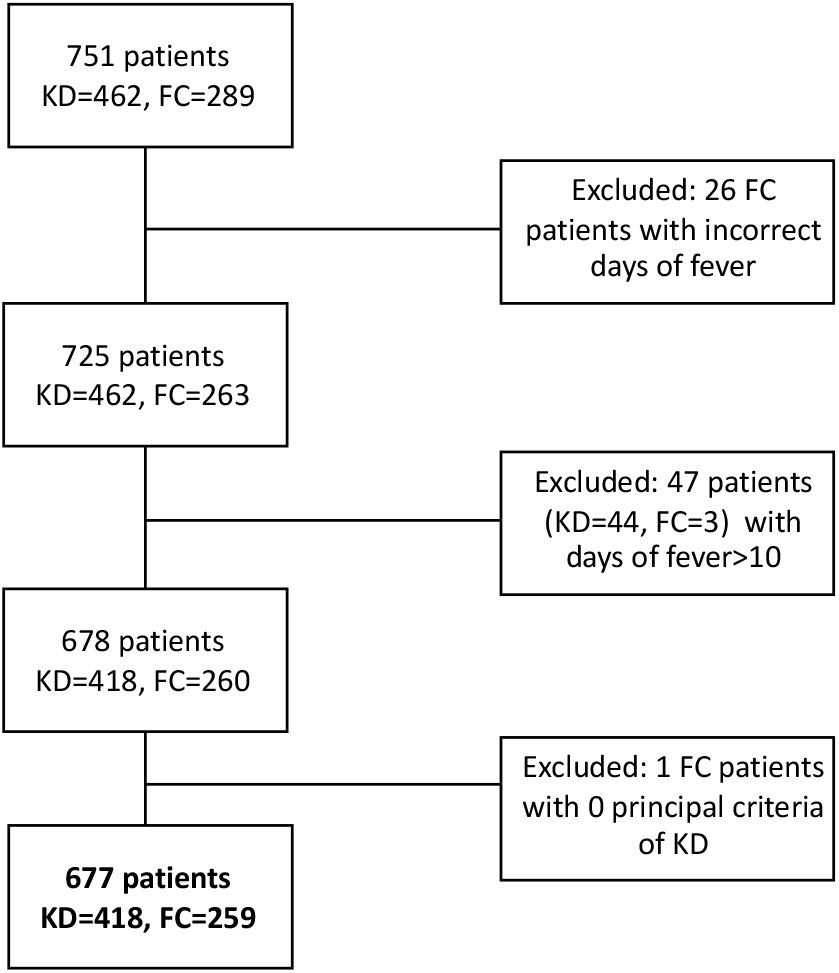
Cohort construction of patients in Chang Gung Cohort

### Univariable analysis of clinical and laboratory test variables

The majority of the patients with KD (368 of 418, 88.0%) had 4 or 5 principal clinical criteria, and 91.3% (217 of 259, 83.7%) of FCs had 1 to 3 principal clinical criteria (Table 3A). Shown in Figure 2, 36 KD and 59 FC patients had three clinical criteria while 14 KD and 135 FC had two clinical criteria. 157/418 KD patients were less than 1 year old while 261/418 KD patients were older than 1 year. Most of the laboratory test variables except C-reactive protein, eosinophils, hemoglobin, and platelet counts differed significantly between patients with KD and FCs in all 3 sub-cohorts (Table 3B).

**Table 3A.**
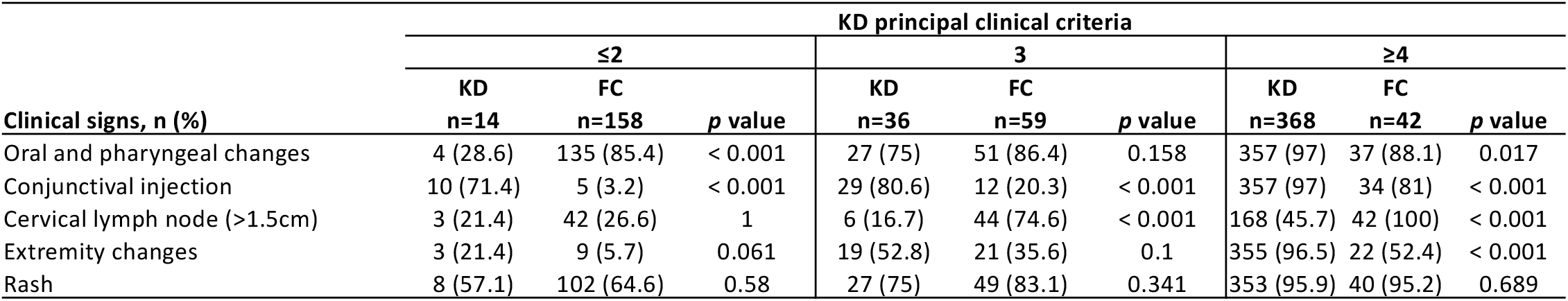
Distribution of the clinical signs among KD and FC patients (count and percentage) in sub cohorts manifesting fewer than 2, 3 or ≥4 principal clinical criteria for KD. *p* value: Fisher’s exact test. KD: Kawasaki disease; FC: febrile control.

| Clinical signs, n (%) | KD principal clinical criteria |  |  |  |  |  |  |  |  |
| --- | --- | --- | --- | --- | --- | --- | --- | --- | --- |
|  | ≤2 |  |  | 3 |  |  | ≥4 |  |  |
|  | KD<br>n=14 | FC<br>n=158 | p value | KD<br>n=36 | FC<br>n=59 | p value | KD<br>n=368 | FC<br>n=42 | p value |
| Oral and pharyngeal changes | 4 (28.6) | 135 (85.4) | < 0.001 | 27 (75) | 51 (86.4) | 0.158 | 357 (97) | 37 (88.1) | 0.017 |
| Conjunctival injection | 10 (71.4) | 5 (3.2) | < 0.001 | 29 (80.6) | 12 (20.3) | < 0.001 | 357 (97) | 34 (81) | < 0.001 |
| Cervical lymph node (>1.5cm) | 3 (21.4) | 42 (26.6) | 1 | 6 (16.7) | 44 (74.6) | < 0.001 | 168 (45.7) | 42 (100) | < 0.001 |
| Extremity changes | 3 (21.4) | 9 (5.7) | 0.061 | 19 (52.8) | 21 (35.6) | 0.1 | 355 (96.5) | 22 (52.4) | < 0.001 |
| Rash | 8 (57.1) | 102 (64.6) | 0.58 | 27 (75) | 49 (83.1) | 0.341 | 353 (95.9) | 40 (95.2) | 0.689 |

**Table 3B.**
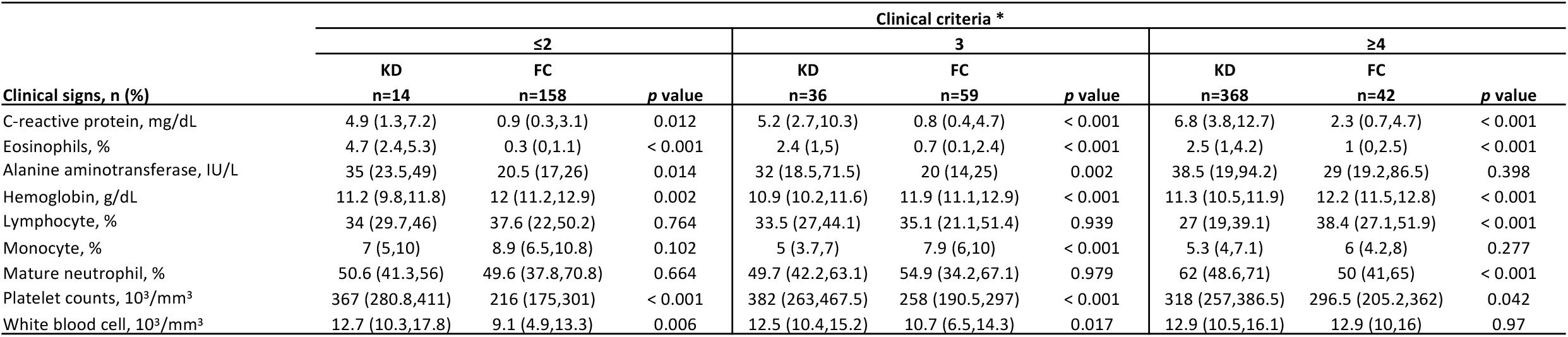
Univariable analysis of laboratory test variables in patients with KD and FCS who had fewer than two, three and at least four clinical criteria

**Figure 2.**
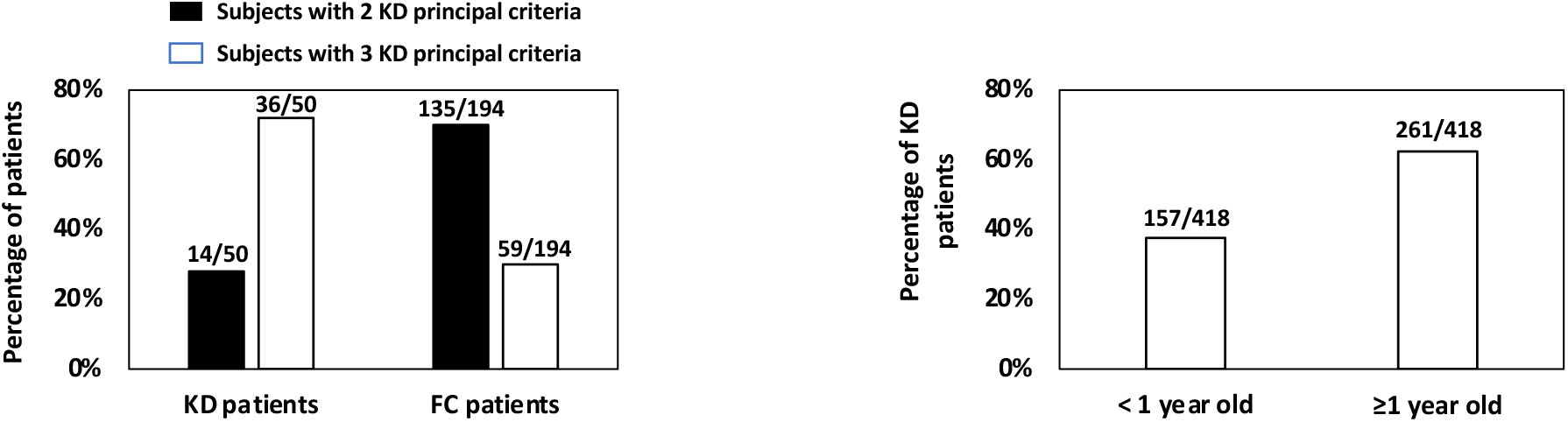
(Left) Percentage of KD and FC patients with 2 or 3 principal clinical KD criteria; (Right) percentage of KD patients with age of either <1 or ≥ 1 year old. FC, febrile control; KD, Kawasaki disease.

### Asia single center blind test results

In this Taiwan single center study (Figure 3), the algorithm’s testing performance is in line with our 2016 US single center [21] and US 2020 multi-center [31] analyses: sensitivity of 90.7% (379/418), specificity of 86.1% (223/259), PPV of 92.7% (379/409), and NPV of 90.3% (223/247); 3.5% (15/418) of the patients with KD and 2.3% (6/259) FCs were classified as indeterminate. Overall, 5.7% (24/418) of the patients with KD and 11.5% (30/259) of FCs were misclassified by the algorithm.

**Figure 3.**
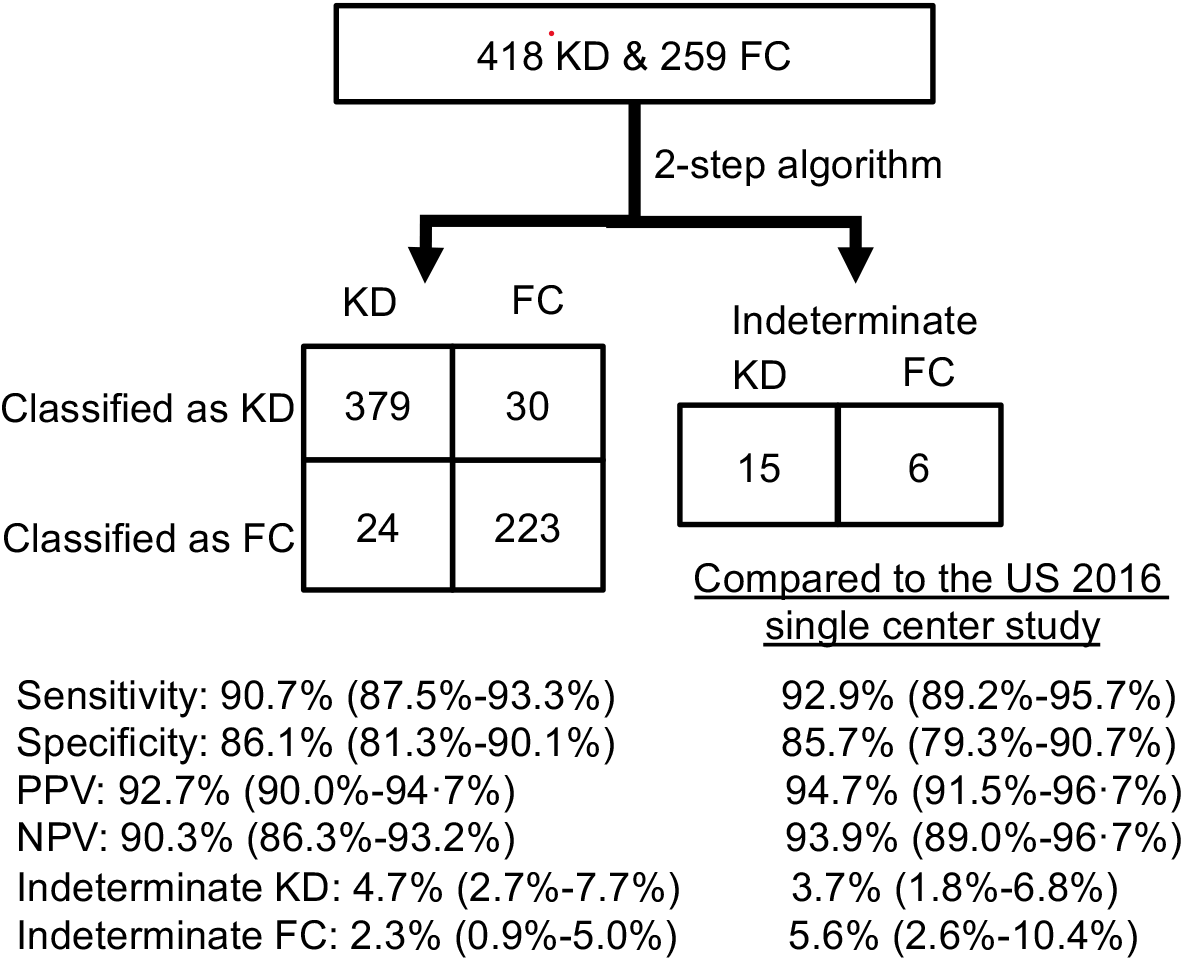
Performance of the two-step algorithm for classification of patients with KD and FCs. Left: a 2×2 table compared with US single-center validation results. The US cohort was described in 2016 Hao et al. publication. Right: percentages of correctly classified, misclassified and indeterminate patients. FCs, febrile controls; KD, Kawasaki disease; NPV, negative positive value; PPV, positive predictive value.

We stratified the patients into < 1 or ≥ 1 year old bins for the performance analysis (Table 4A, top panel). Of the 157/418 KD infants younger than 1 year, 89.6% were classified correctly, and 7.0% were indeterminate. Of patients with KD ≥ 1 year old, 90.3% were classified correctly, and 8.0% were indeterminate. Shown in Figure 4, for the 379 correctly classified KD, 140/379 were less than 1 year old and 239/379 were older than 1 year old. For the 24 KD wrongly classified into FC, 10/24 were less than 1 year old and 14/24 were older than 1 year old. For the 15 KD wrongly classified into intermediate, 7/15 were less than 1 year old and 8/15 were older than 1 year old.

**Table 4A.**
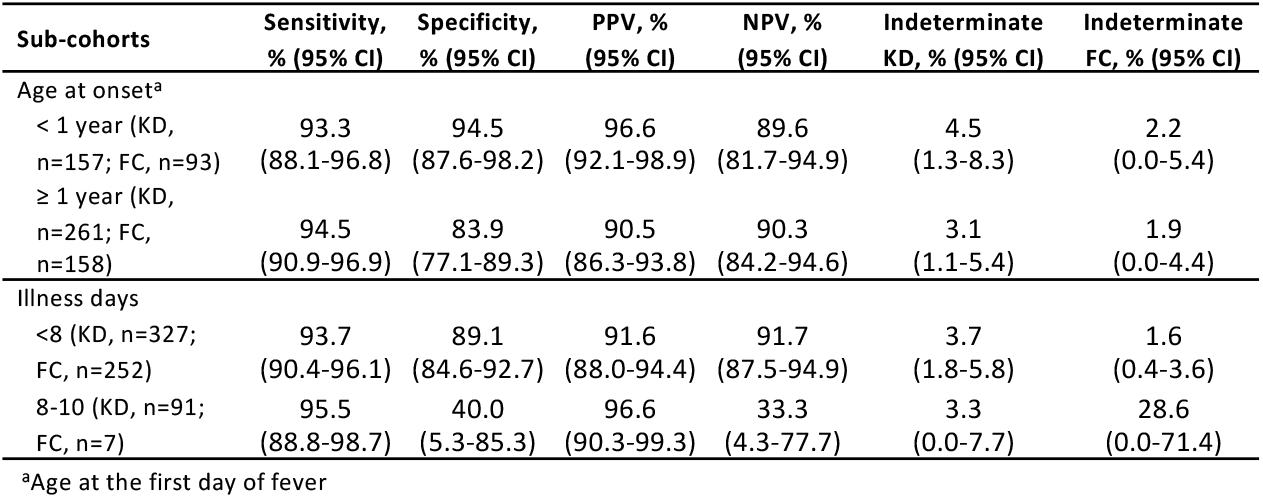
Performance of the 2-step algorithm in Kawasaki disease patients and febrile controls stratified by age at onset and days of illness. There were 8 FC subjects missing age information at the first day of fever..

**Figure 4.**
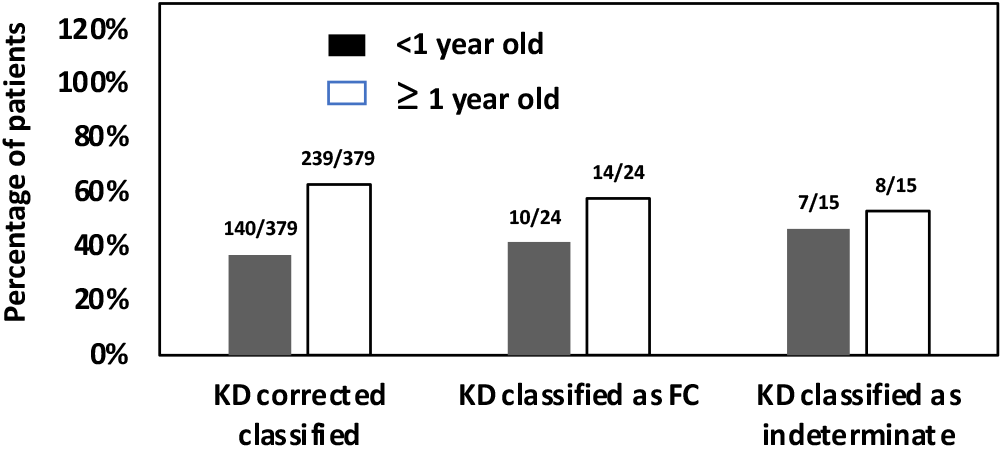
Percentage of KD patients, (left) correctly classified, (middle) wrongly classified as FC, (right) wrongly classified as indeterminate, stratified by age of less than or greater than 1 year. FC, febrile control; KD, Kawasaki disease.

We stratified the patients into < 8 or 8-10 illness day bins for the performance analysis (Table 4B, bottom panel). Of the 327/418 KD patients with < 8 illness days, 93.7% were classified correctly, and 3.7% were indeterminate. Of the 91/418 KD patients with 8-10 illness days, 95.5% were classified correctly, and 3.3% were indeterminate. Of the 252/259 FC patients with < 8 illness days, 89.1% were classified correctly, and 1.6% were indeterminate. Of the 7/259 FC patients with 8-10 illness days, 40.0% were classified correctly, and 28.6% were indeterminate. The poor performance, as previously observed in US multi-center study [31], the specificity (40.0%) and NPV (33.3%), of 8-10 illness day FCs may be due to the small sample size (n=7 FCs with 8-10 illness days). We also compared, the ratio of median laboratory test values between the KD and FC patients (Table 4B), of the bins of illness day less than 8 or illness day 8-10. Much more up regulated ratios, of the tests of C-reactive protein, Alanine aminotransferase and while blood cell, were observed when contrasting between the bins of illness day less than 8 and illness day 8-10 (*p* value < 0.01).

**Table 4B.** Comparison of ratio of median laboratory values in Kawasaki disease patients to that in febrile controls.

| Variable | Ratio of median in Kawasaki disease patients to median in febrile controls |  |
| --- | --- | --- |
|  | Illness days <8 (KD, n=327; FC, n=252) | Illness days 8-10 (KD, n=91; FC, n=7) |
| C-reactive protein | 3.022 | 1.003 |
| Eosinophils | 2.794 | 3.105 |
| Alanine aminotransferase | 2.659 | 0.82 |
| Hemoglobin | 0.92 | 0.933 |
| Lymphocyte | 0.825 | 0.872 |
| Monocyte | 0.698 | 1.001 |
| Mature neutrophil | 1.133 | 1.022 |
| Platelet counts | 1.299 | 1.127 |
| White blood cell | 1.234 | 0.976 |

There were 50/418 KD patients who presented with 3 or fewer criteria and 368/418 KDs with 4 or 5 clinical criteria (Figure 5). Compared with its sensitivity in KD patients with 4 or 5 clinical criteria (361/368, 98.0%), the algorithm had a lower sensitivity for KD with less than or equal to 2 criteria 14.2% (2/14) and for KD with 3 criteria 44.4% (16/36) (Fisher’s exact test *p* < 0·0001). Compared with its specificity in FC patients with 4 or 5 clinical criteria (10/42, 23.8%), the algorithm had a higher specificity for FC with less than or equal to 2 criteria 99.4% (157/158) and for FC with 3 criteria 94.9% (56/59) (Fisher’s exact test *p* < 0·0001). For the total of 15 KDs and 6 FCs wrongly classified into intermediate, 1/13/1 KDs and 1/3/2 FCs were from the bins of with less than or equal to 2, with 3, or with greater than 4 clinical criteria, respectively.

**Figure 5.**
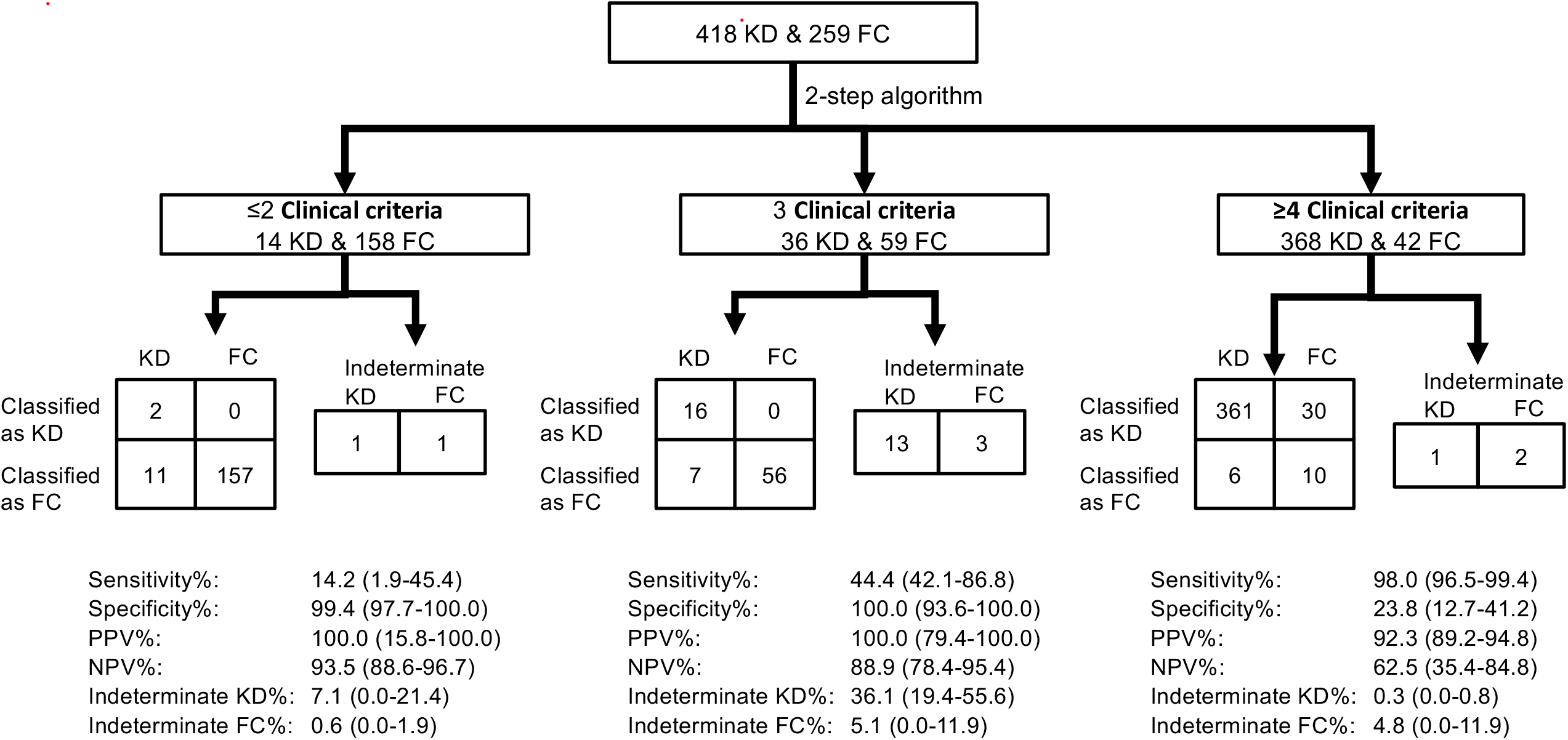
. Performance of the two-step algorithm for classification of patients with KD and FCs stratified by number of KD principal clinical criteria. FCs, febrile controls; KD, Kawasaki disease; NPV, negative positive value; PPV, positive predictive value. A. 2×2 tables; B. Predictive performance comparison.

### Classification of KD patients with coronary artery abnormalities

Coronary artery abnormalities were documented in 44 of the 418 KD patients (10.5%) (Table 5). One patient was not classified correctly. The misclassified KD patient had a maximum Z-score of 2·8 that resolved by 8 weeks after treatment. This is still a problem, if we use the criteria for treatment. All 8 KDs with aneurysm were identified by the two-step algorithm.

**Table 5.** Performance of the two-step algorithm in relation to echocardiogram results.

| Coronary artery status by echocardiogram, n | KD, correctly classified | KD, classified indeterminate | KD, misclassified |
| --- | --- | --- | --- |
| **Normal**<br>(RCA&LAD always <2.5) | 309 | 10 | 12 |
| **Dilated**<br>(Z score (RCA or LAD) $\geq 2.5$, and resolve in 8 weeks) | 30 | 0 | 1 |
| **Aneurysm**<br>(Z score $\geq 5$, or dilated segment >1.5) | 8 | 0 | 0 |
| **Others** |  |  |  |
| Z score (RCA or LAD) $\geq 2.5$, but not resolve in 8 weeks | 12 | 0 | 0 |
| Z score (RCA or LAD) $\geq 2.5$, but had no data after 8 weeks | 1 | 0 | 0 |
| Data missing | 10 | 0 | 0 |
| No 2D echo | 9 | 5 | 11 |

\* RCA: Right coronary artery; LAD: Left anterior descending. 2D echocardiography: 2D echo.

### Comparison of clinical and laboratory data between Asian single-center and US 2016 single-center [21] and 2020 multi-center [31] studies

The frequencies of the five KD principal criteria and illness days in this Taiwan single-center study were in line with those in the US single-center **[21]** and multi-center **[31]** validation dataset of the algorithm except for cervical lymph node (25.5%/19.5% versus 49.4%), conjunctival injection (44.1%/58.9% versus 19.7%), oropharyngeal changes (48.4%/46.8% versus 86.1%), and days of illness (5.0/5.0 versus 2.0) in FC subjects (Table 6).

**Table 6.**
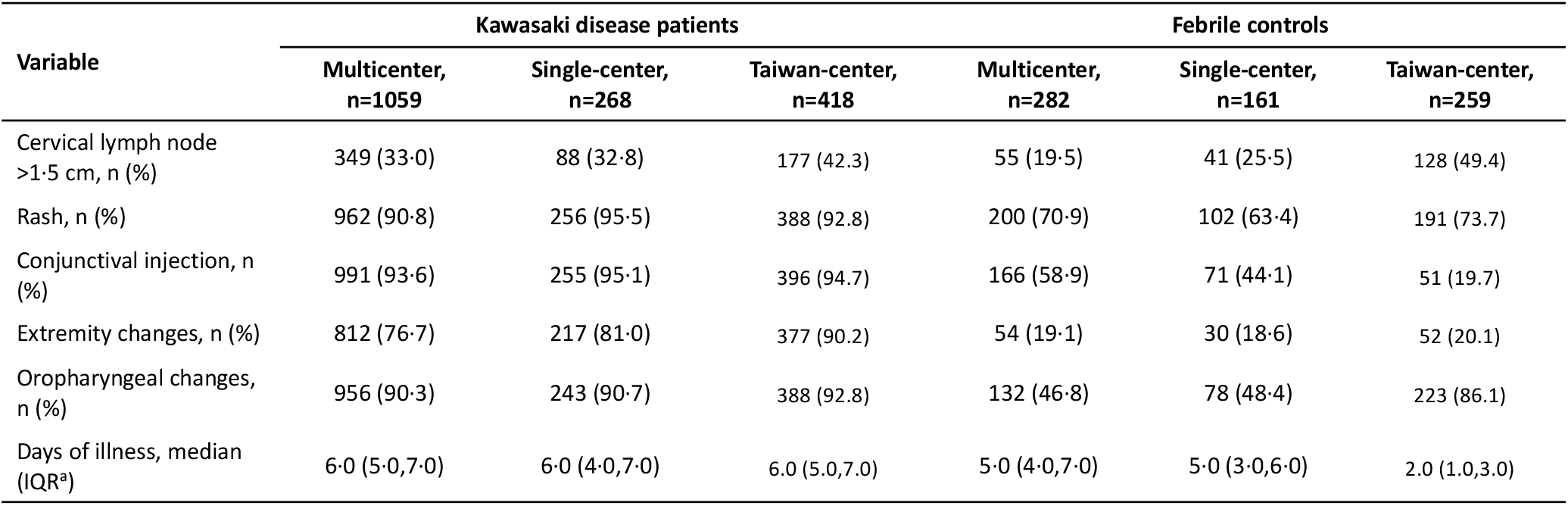
Clinical variables of Kawasaki disease patients and febrile controls in the 2020 US multicenter, 2016 US single-center and current Asia single-center validation studies.

| Variable | Kawasaki disease patients |  |  | Febrile controls |  |  |
| --- | --- | --- | --- | --- | --- | --- |
|  | Multicenter,<br>n=1059 | Single-center,<br>n=268 | Taiwan-center,<br>n=418 | Multicenter,<br>n=282 | Single-center,<br>n=161 | Taiwan-center,<br>n=259 |
| Cervical lymph node<br>>1.5 cm, n (%) | 349 (33.0) | 88 (32.8) | 177 (42.3) | 55 (19.5) | 41 (25.5) | 128 (49.4) |
| Rash, n (%) | 962 (90.8) | 256 (95.5) | 388 (92.8) | 200 (70.9) | 102 (63.4) | 191 (73.7) |
| Conjunctival injection, n<br>(%) | 991 (93.6) | 255 (95.1) | 396 (94.7) | 166 (58.9) | 71 (44.1) | 51 (19.7) |
| Extremity changes, n (%) | 812 (76.7) | 217 (81.0) | 377 (90.2) | 54 (19.1) | 30 (18.6) | 52 (20.1) |
| Oropharyngeal changes,<br>n (%) | 956 (90.3) | 243 (90.7) | 388 (92.8) | 132 (46.8) | 78 (48.4) | 223 (86.1) |
| Days of illness, median<br>(IQR<sup>a</sup>) | 6.0 (5.0,7.0) | 6.0 (4.0,7.0) | 6.0 (5.0,7.0) | 5.0 (4.0,7.0) | 5.0 (3.0,6.0) | 2.0 (1.0,3.0) |

## DISCUSSION

It is essential to diagnose KD early in the acute phase as early administration with anti-inflammatory therapies can reduce the risk of developing coronary artery aneurysms [1]. However, the diagnosis of KD remains difficult as the clinical signs overlap with those of other pediatric febrile illnesses. In this study, we tested a US multi-center validated KD diagnostic algorithm using a blinded Asian single-center cohort. The algorithm had a sensitivity of 90.7%, a specificity of 86.1%, a PPV of 92.7%, and a NPV of 90.3%, similar to that of the US single-center and multi-center validation studies [10, 21, 31]. The algorithm identified 43/44 (98.0%) of patients with KD who developed coronary artery abnormalities, and only misclassified as one FC patient with Z score (RCA or LAD) ≥2.5, and resolve in 8 weeks. In this study, all 8 KD subjects with aneurysms were correctly diagnosed by the 2-step algorithm.

Given that this algorithm has been validated in US, this blinded Asian single center study shall provide evidence to support a multi-center prospective testing in an Asian setting: where the KD prevalence should be higher than US; and in which KD patients are rare and febrile controls are common to evaluate its potential utility as a physician support tool.

We recognize both strengths and weaknesses in this study. The strength of the study is the demonstration of the robustness and effectiveness of the algorithm and the algorithm portability from North America to Asia. The KD and FC subjects in the Asian single-center are generally younger than the US single-center and multi-center analyses, thus demonstrating its potential clinical utility across a broad range of similar appearing illnesses. However, in this Asian study, the clinical parameters of bands, GGT, and ESR were not routinely collected in the Taiwan Chang Gung Hospital for the KD patients, therefore, not used in the two-step algorithm. If collected in the future, the algorithm performance might be improved when prediction with Asian patients. The FCs were not examined by the echocardiography, which may limit our ability to completely rule out KD diagnosis. In this Asian single-center study, we had a larger number of KD patients than FCs as in the US multi-center study, and the frequency of KD patients in a prospective study in the emergency department setting will be much lower, which will undoubtedly worsen the performance of this algorithm. The actual ratio of KD to FC patients who would be screened for KD is not known and shall warrant more studies. We are planning a multi-center study in the Asian emergency room setting to assess the prevalence of KD and prospectively evaluate the utility of this algorithm.

## CONCLUSIONS

We assessed the performance of our 2-step algorithm in a blinded, single-center study in Asia. This work demonstrates the algorithmic portability from North America to Asia, providing further support for moving forward with an adequately powered, multicenter study in the Asian emergency department settings to assess if this algorithm will be a useful clinical support tool to manage Kawasaki disease in Asia where the disease prevalence is higher than in US.

## Data Availability

All data produced in the present study are available upon reasonable request to the corresponding authors

## Acknowledgement

This research was funded by the following grants CPRPG8H0051-2, CMRPG8L0011 and CMRPG8L1241 provided by Chang Gung Memorial Hospital.

## List of abbreviations

KD: Kawasaki disease
IVIG: Intravenous Immunoglobulin
AHA: American Heart Association
LDA: Linear discriminant analysis
WBC: White blood cell
CRP: C-reactive protein
GGT: Gamma glutamyl transferase
RCA: Right coronary artery
LAD: Left anterior descending
PPV: Positive predictive value
NPV: Negative positive value

## What is already known on this topic

A key objective of research in KD is to reduce the risk of long-term cardiovascular sequelae by expediting timely diagnosis.

The major challenge in diagnosing KD is that it shares clinical signs with other childhood febrile illnesses.

A computer-based algorithm was validated using clinical criteria and laboratory tests to differentiate US single-center and multi-center patients with KD from those with other febrile illnesses.

## What this study adds

This is the first blinded, single-center testing of a US validated computer algorithm for the differentiation of patients with KD from others with clinically similar febrile illnesses in Asia.

The algorithm has sufficient sensitivity and PPV to identify the majority of patients with KD diagnosed at either single center in Asia or centers across North America.

The results suggest a promising approach to the timely diagnosis and treatment of KD patients to reduce the risk of coronary artery aneurysms. The results also provide evidence to support the launch of the multi-center validation trial in Asia.

