## Supplementary table for "Single center blind testing of a US multi-center validated diagnostic algorithm for Kawasaki disease in Asia"

| Variable | Weight in 2-step algorithm |
|---|---|
| Days of illness | 0.108 |
| Rash | 0.882 |
| Conjunctival injection | 1.152 |
| Oropharyngeal changes | 1.046 |
| Cervical lymph node >1.5 cm | 0.350 |
| Extremity changes | 1.291 |
| White blood cell count | 0.030 |
| Neutrophils, % | 0.0009 |
| Lymphocytes, % | -0.009 |
| Monocytes, % | -0.013 |
| Eosinophils, % | 0.032 |
| Hemoglobin concentration | -0.229 |
| C-reactive protein | 0.021 |
| Platelet count | 0.001 |
| Alanine aminotransferase | 0.002 |

Supplementary Table 1A. Predictive variables and their corresponding weights in the LDA algorithm.

| Variable | 2 clinical criteria | 3 clinical criteria | 4 or 5 clinical criteria |
|---|---|---|---|
| Days of illness | 1.00 | 6.95 | 3.60 |
| Rash | 0.00 | -1.84 | 0.00 |
| Conjunctival injection | 1.40 | -0.48 | -1.27 |
| Oropharyngeal changes | -1.90 | -1.72 | 1.00 |
| Cervical lymph node >1.5 cm | -0.02 | 1.58 | 0.20 |
| Extremity changes | -1.37 | 3.81 | -0.94 |
| White blood cell count | 8.87 | 1.71 | 5.42 |
| Neutrophils, % | 7.34 | 2.82 | -0.06 |
| Lymphocytes, % | 0.28 | 0.25 | 6.48 |
| Monocytes, % | 0.57 | 0.87 | 0.55 |
| Eosinophils, % | 2.13 | 3.47 | -2.09 |
| Hemoglobin concentration | 0.18 | 13.90 | 6.36 |
| C-reactive protein | 7.72 | 8.07 | 5.38 |
| Platelet count | 2.01 | 8.18 | 3.70 |
| Alanine aminotransferase | -1.68 | 3.34 | 8.56 |

Supplementary Table 1B. Predictive variables and their corresponding weights in the random forest algorithm.
